# Epigenetic pathways and risk factors for Type 2 Diabetes in a Middle Eastern Cohort

**DOI:** 10.1101/2025.04.21.25326110

**Authors:** Noha A Yousri, Yanwei Cai, Boya Guo, Gina M Peloso, Ida Surakka, Ulrike Peters, Heather M. Highland, Nady El Hajj, Burcu F. Darst, Misa Graff, Rayaz A Malik

## Abstract

Type 2 diabetes (T2D) is a complex metabolic disorder influenced by genetic, epigenetic, and environmental factors. Triglycerides and lipoproteins play a role in the development and progression of both macro and micro vascular complications of diabetes and our previous studies demonstrated a correlation between DNA methylation of genes associated with diabetes and diabetes metabolic pathways. We now investigate the association of DNA methylation in diabetes with alterations in lipoproteins and triglycerides and genomic risk scores (GRS), in subjects from the Qatar Biobank population. We have identified *TXNIP* and *PFKFB2* gene methylation to have a mechanistic link with altered lipoproteins and triglycerides in T2D.

## Introduction

Type 2 diabetes (T2D) is a multifactorial disease characterized by insulin resistance and β-cell dysfunction, with epigenetic alterations, in particular DNA methylation, which play critical roles in its pathogenesis [Yousri et al, 2023]. Genetic predisposition and lifestyle factors contribute to altered methylation in T2D. Diabetes-related complications may also arise due to DNA methylation or, conversely, contribute to epigenetic modifications, creating a complex feedback system.

Dyslipidemia is a well-established feature of T2D, leading to an abnormality in HDL-C, LDL-C, triglycerides, and total cholesterol levels. However, it remains unclear whether these lipid abnormalities are a cause or consequence of diabetes [Haase C. L., et al, 2015, Zhu Z., et al, 2022]. Our previous studies revealed that diabetes-related metabolic pathways were associated with metabolic syndrome, retinopathy and dyslipidemia [Yousri at el,2022] and we also identified 66 CpG sites whose methylation sites associated with T2D [Yousri et al, 2023]. Some of these methylated CpG sites were associated with certain metabolites that we have identified to be correlated with lipid traits, for example, the multi-omics network identified an alanine subnetwork that links *PFKFB2* and *TXNIP* to the urea cycle and to amino acids, where 1-carboxyethylphenylalanine was shown to have a significant association with triglycerides.

In this study, we aim to refine our understanding of how altered lipoprotein and triglyceride levels interact with epigenetic and genomic factors in T2D. By leveraging methylation profiles, whole-genome sequencing, lipoprotein and triglyceride data from the Qatar BioBank, we have investigated the relationship between these risk factors and DNA methylation signatures associated with T2D.

## Methods

A total of 1000 samples, from the Qatar Bio Bank [Al Thani, A, et al, 2019], with methylation profiles, whole genomes, T2D status, lipoproteins, triglyceride, age, BMI, HbA1c, gender and cell counts were used for the analysis. We focused on the 66 CpG methylation sites associated with T2D status as previously described [Yousri et al, 2023] and tested for their association with 1) lipid traits: HDL-C, LDL-C, TG, TC and non-HDL, 2) the genomic risk score (GRS) of T2D, and 3) GRS of the aforementioned lipid traits. Genomic risk scores (GRS) for T2D were calculated using a validated score from [Ge et al,2022]. The GRS for the lipid traits were computed using published results from our previous work on multi-ancestry GLGC GWAS [Graham et al. 2021], including 1,663,048 individuals, after subtracting validation samples used to test the scores. GRS weights were derived using the same pipeline and parameters as [Graham et al. 2021], applying both the PT method [ISC et al, 2009] (M1) and PRS-CS method [Ge et al,2022] (M2). GRSs were validated in the Qatari cohort by computing associations of the GRS with their respective trait in a larger set of samples with whole genomes. Supplementary Tables (ST1-ST11) indicate the sample size used in each analysis, and in T2D and controls separately, adjusting for relevant covariates.

LDL-C and TC levels were adjusted for use of lipids lowering medication such as statins; the pre-medication levels were approximated by dividing the original LDL-C values by 0.7 and the original TC value by 0.8). Triglycerides values were log transformed. We removed outliers (more than six standard deviations), and normalized values by rank-based inverse-normal transformation. Statistics describing the traits are in Supplementary Table ST12.

For validation of GRS in all traits including T2D, residuals of the trait were obtained from kinship correction then were corrected for all covariates and inverse normalized and the resulting residuals used for analysis. The GRS values were z-scored for normalization before regression with the traits.

Linear regression models were used to assess associations of 66 T2D CpGs with lipoproteins, triglycerides and the GRSs, correcting for gender, cell counts, batch effects, smoking surrogate (AHRR), genomic principal components (PC1 to PC3), well position, and plate. We excluded age and BMI due to multi-collinearity (Supplementary Table ST13) with the traits and T2D. The CpGs have previously been checked for their association with age [Yousri et al, 2023] and two models have been adopted, with and without BMI. Statistical significance was defined using a Bonferroni adjustment accounting for 66 CpG sites with an alpha of 0.05.

## Results

We aim to identify the effect of lipoproteins and triglycerides on the epigenetic mechanisms of T2D using the 66 CpG sites associated with T2D [Yousri et al, 2023], and the GRS scores of T2D and lipoprotein and triglycerides, computed in our cohort using previously published GWAS results. To identify such mechanistic links, we performed the following analyses: 1) Computed associations of the 66 T2D CpGs with the levels of lipoproteins (LDL-C, HDL-C, non-HDL-C total cholesterol (TC)), and triglycerides, and 2) Computed associations between the 66 CpGs and genomic risk scores (GRS) of T2D, and all aforementioned traits. Figure 1 summarizes the study. shows the correlation of T2D with its GRS and the GRSs of the selected traits. A total of 13 significant associations were identified between the 66 T2D CpG sites and lipid traits, including 4 unique CpG sites located in *TXNIP* and *PFKFB2* (Bonferroni adjusted p < 7.5 x 10^-4^) (Table 1). GRS scores were regressed against their respective traits, and in stratified T2D and control samples (Table 2). The T2D GRS showed significant associations with the lipid traits and their GRSs (Figure 2 and Table 2). CpG sites in *TXNIP* showed Bonferroni significant associations with the GRSs of T2D, log TG and HDL-C, and CpG sites in *PFKFB2* was nominally significant associated with T2D GRS (Table 3).

**Table 1.** T2D CpGs associations with lipoproteins LDL-C, HDL-C, triglycerides (log TG), non-HDL-C and total cholesterol (TC).

| CpG | Gene | N | Beta | SE | pval | trait |
| --- | --- | --- | --- | --- | --- | --- |
| cg19693031 | *TXNIP* | 999 | -0.0156592 | 0.00186418 | **1.52E-16** | log_TG |
| cg02988288 | *TXNIP;NBPF20;NBPF10* | 999 | -0.0081794 | 0.0011685 | **4.69E-12** | log_TG |
| cg19693031 | *TXNIP* | 998 | -0.0099421 | 0.00144996 | **1.23E-11** | Non-HDL |
| cg19693031 | *TXNIP* | 998 | -0.0090747 | 0.00182766 | **8.07E-07** | TC |
| cg02988288 | *TXNIP;NBPF20;NBPF10* | 998 | -0.0044924 | 0.00091499 | **1.06E-06** | Non-HDL |
| cg22699725 | *PFKFB2* | 999 | -0.0074639 | 0.00156702 | **2.19E-06** | HDL |
| cg22699725 | *PFKFB2* | 999 | 0.00639117 | 0.0014381 | **9.81E-06** | log_TG |
| cg22699725 | *PFKFB2* | 998 | 0.00464413 | 0.00111338 | **3.29E-05** | Non-HDL |
| cg00683922 | *PFKFB2* | 999 | 0.00647244 | 0.00155993 | **3.62E-05** | log_TG |
| cg19693031 | *TXNIP* | 999 | 0.00814248 | 0.00208901 | **1.04E-04** | HDL |
| cg19693031 | *TXNIP* | 988 | -0.0068227 | 0.00182411 | **1.94E-04** | LDL |
| cg02988288 | *TXNIP;NBPF20;NBPF10* | 998 | -0.0040594 | 0.00114745 | **4.22E-04** | TC |
| cg00683922 | PFKFB2 | 998 | 0.00410353 | 0.00120725 | **7.03E-04** | Non-HDL |
| cg00683922 | PFKFB2 | 999 | -0.0056866 | 0.00170737 | 8.98E-04 | HDL |
| cg00929203 |  | 999 | -0.0031427 | 0.00105098 | 2.86E-03 | log_TG |
| cg26747273 | IDO2 | 999 | -0.0063072 | 0.00217291 | 3.78E-03 | log_TG |
| cg02988288 | TXNIP;NBPF20;NBPF10 | 999 | 0.00374356 | 0.0013006 | 4.08E-03 | HDL |
| cg06555354 | SPRED2 | 999 | -0.0047335 | 0.00166029 | 4.45E-03 | log_TG |
| cg27094813 | -- | 999 | -0.0036531 | 0.00135598 | 7.18E-03 | log_TG |
| cg09879165 | CACNA2D2 | 998 | -0.0023069 | 0.00088672 | 9.42E-03 | TC |
| cg21124952 | -- | 999 | -0.0035655 | 0.00137376 | 9.58E-03 | log_TG |
| cg27094813 | -- | 998 | -0.0033892 | 0.00130661 | 9.63E-03 | TC |
| cg02988288 | TXNIP;NBPF20;NBPF10 | 988 | -0.002985 | 0.00116164 | 1.03E-02 | LDL |
| cg09879165 | CACNA2D2 | 988 | -0.0021938 | 0.00089394 | 1.43E-02 | LDL |
| cg26747273 | IDO2 | 998 | -0.0040811 | 0.00168336 | 1.55E-02 | Non-HDL |
| cg09879165 | CACNA2D2 | 999 | -0.0021571 | 0.00091964 | 1.92E-02 | log_TG |
| cg00929203 |  | 998 | -0.0017777 | 0.00081327 | 2.91E-02 | Non-HDL |
| cg08992189 | HK1 | 999 | -0.0035565 | 0.00163432 | 2.98E-02 | log_TG |
| cg21124952 |  | 998 | -0.0026901 | 0.0013271 | 4.29E-02 | TC |
| cg26747273 | IDO2 | 998 | -0.0042211 | 0.00210096 | 4.48E-02 | TC |

**Table 2.**
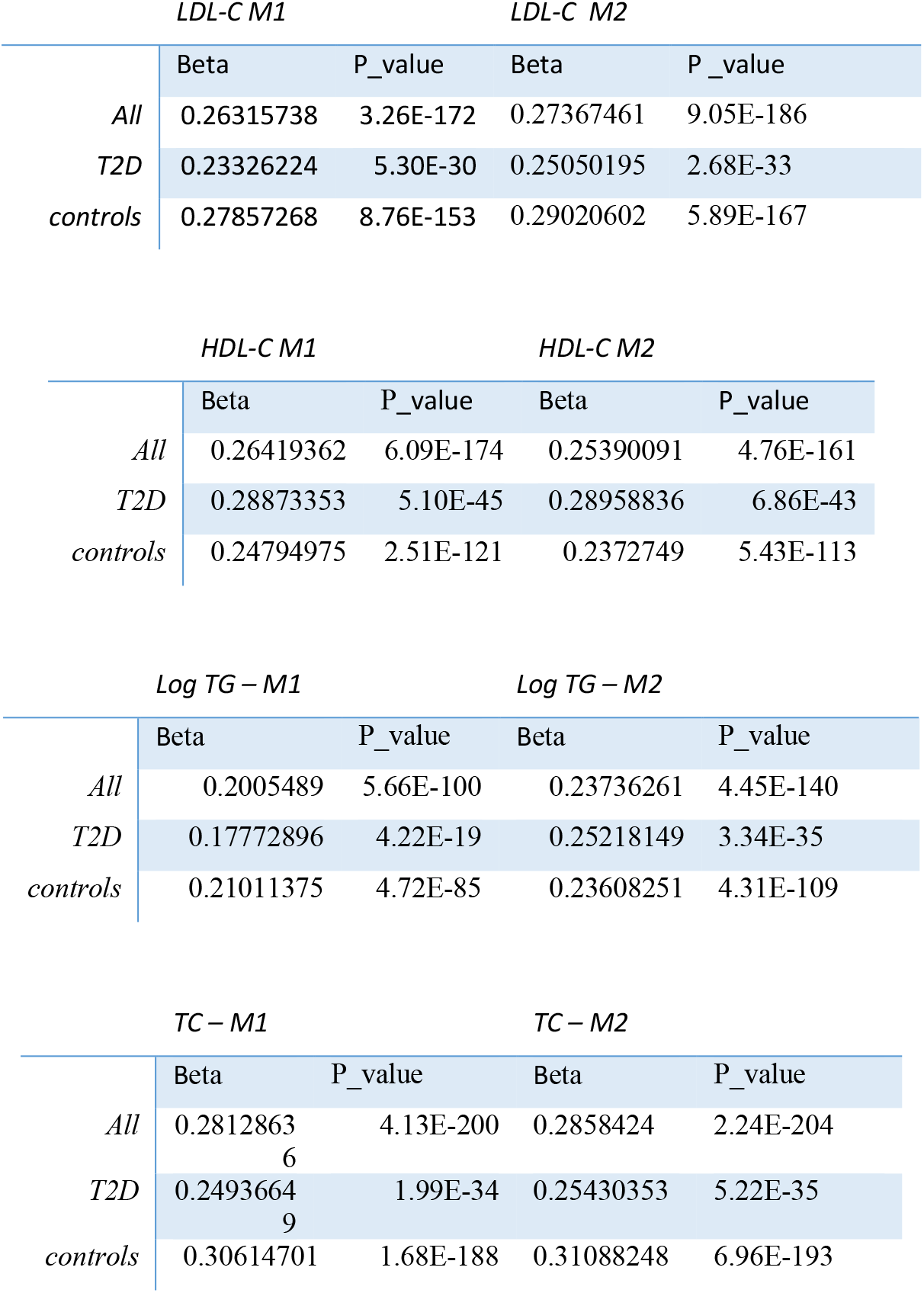
Associations of GRSs of LDL-C, HDL-C, TG and TC with their levels in all samples vs their levels in stratified T2D and controls samples.^2^

**Table 3.**
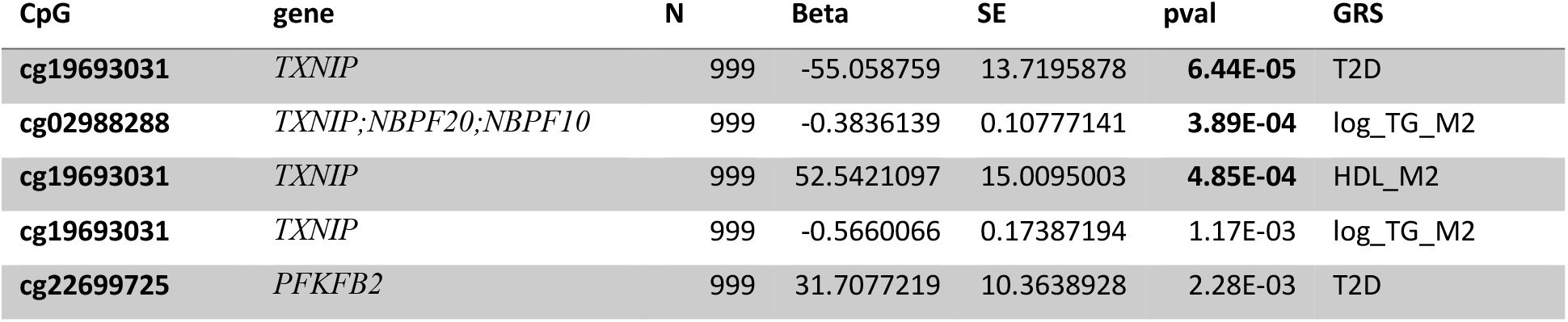
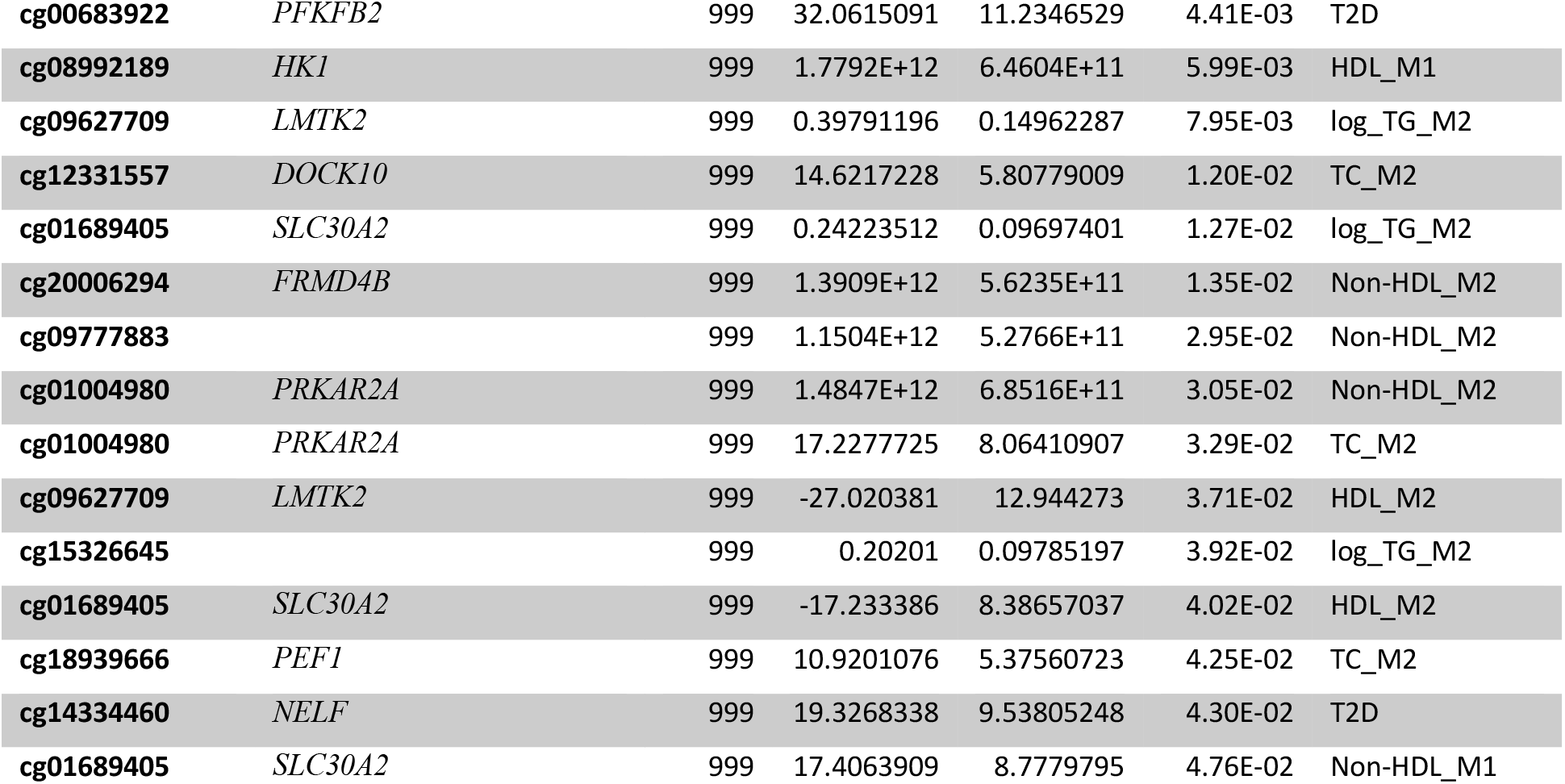
Associations of T2D CpGs with genomic risk scores for T2D, lipoproteins LDL-C, HDL-C, triglycerides (log TG), non-HDL-C and total cholesterol (TC).

**Figure 1.**
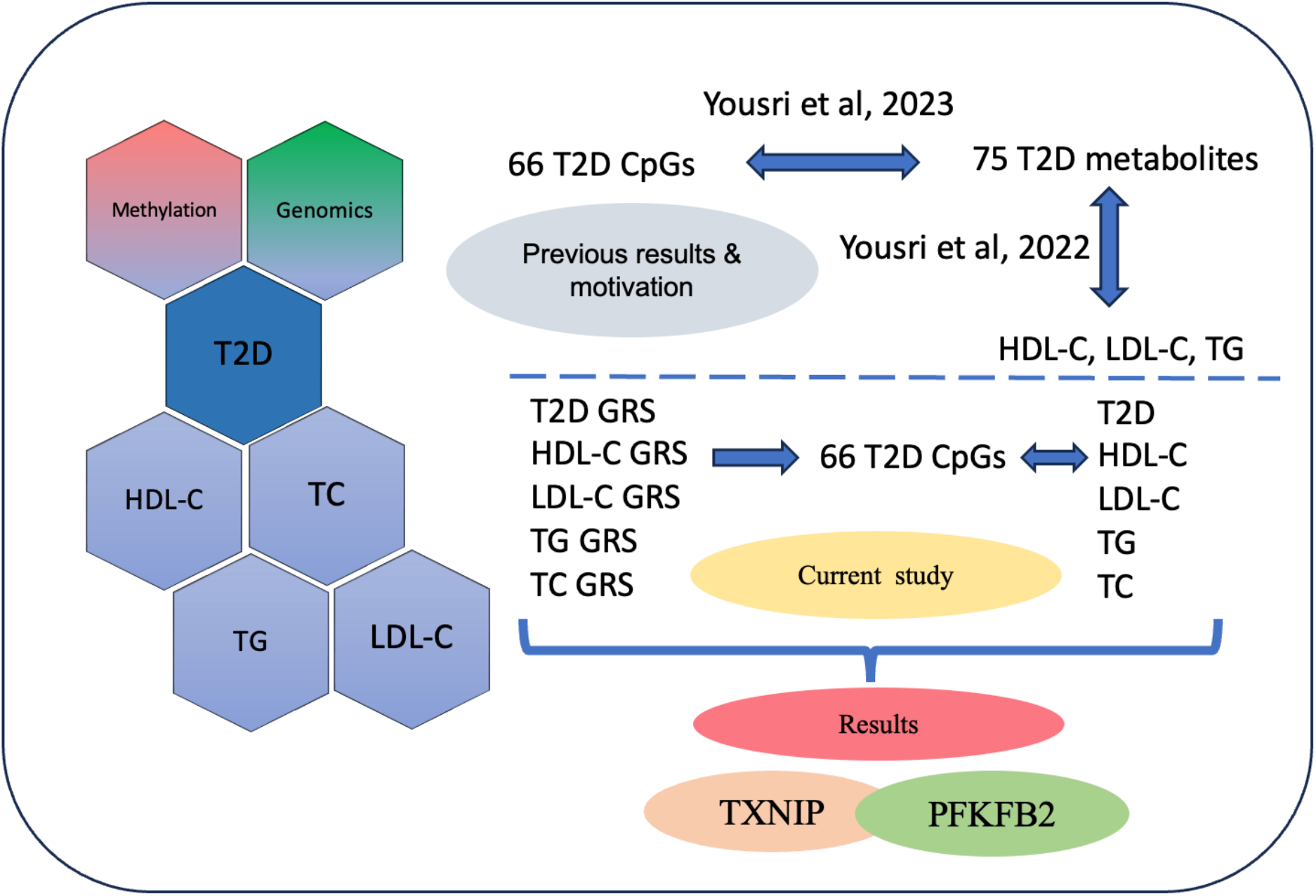
Summary of the main motivation, objectives and results of the study

**Figure 2.**
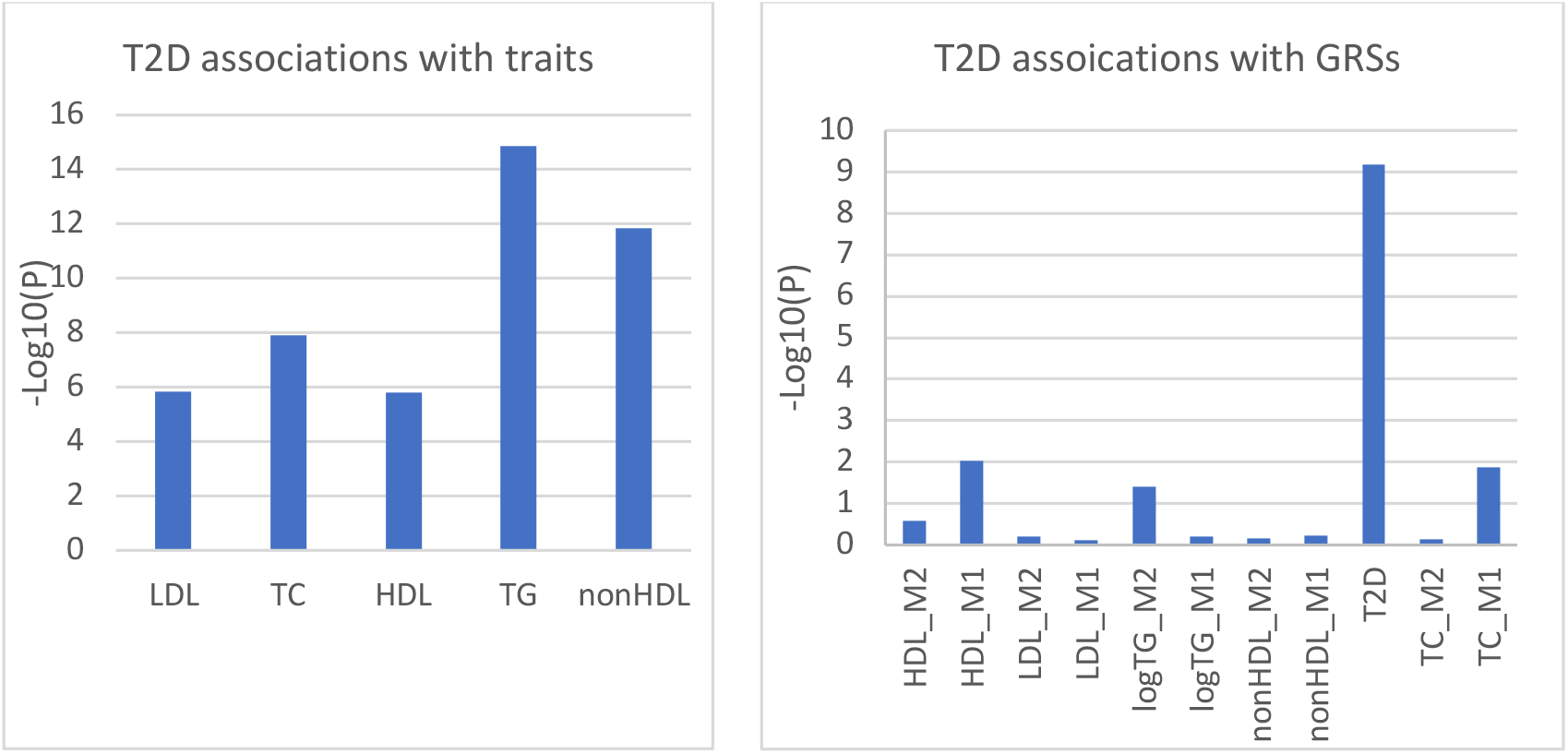
Regression results of T2D with lipoproteins and triglycerides (left) and GRSs (right).

## Discussion

Based on the multi-omics network identified in our previous study [Yousri et al, 2023], we have explored the association of T2D CpG sites with diabetes risk factors. We [Yousri et al, 2022] have previously shown that metabolic networks that combine triglycerides with T2D metabolic pathways share pathways between T2D and lipids. The multi-omics network of metabolites and T2D CpG sites revealed a number of metabolites associated with HDL-C, LDL-C, BMI and triglycerides (Supplementary Table ST14), confirming links between T2D methylated genes and metabolic pathways of triglycerides and lipoproteins. This study revealed that *TXNIP* (Thioredoxin-interacting protein) and *PFKFB2* (Phosphofructokinase-2 isoform B), have important roles in mechanistic pathways linking T2D with HDL-C, LDL-C, TG and total cholesterol. Both genes were central in the multi-omics network study by [Yousri et al, 2023], and were found to be associated with metabolites associated with triglycerides and lipoproteins. *TXNIP* methylation is associated with reduced β-cell dysfunction and lower T2D risk [Juvinao-Quintero, D.L., 2021], [Yousri et al, 2023], and have been reported in almost all studies on T2D methylation. Genetic factors driving *TXNIP* and *PFKFB2* methylation were previously identified in SLC2A1 (GLUT1) and *PFKFB2* [Yousri et al, 2023] and a causal pathway of *PFKFB2* with HbA1c was identified in the same study. Targeting *TXNIP* and *PFKFB2* methylation could be a novel therapeutic strategy for metabolic disorders associated with T2D.

*TXNIP* is a protein that interacts with the thioredoxin system and has been implicated in the regulation of oxidative stress, apoptosis, and glucose homeostasis [Choi et al, 2023]. It acts as a negative regulator of the thioredoxin system, leading to increased oxidative stress. *TXNIP* expression was upregulated in kidneys from a diabetic mouse model, as well as mesangial cells exposed to high glucose, changes of which were associated with specific histone modifications [De Marinis, et, al,]. It is an important regulator of glucose and lipid metabolism through regulation of β-cell function, hepatic glucose production, peripheral glucose uptake, adipogenesis, and substrate utilization [Alhawiti et al, 2017]. Furthermore, *TXNIP* has been shown to be upregulated in insulin-resistant tissues, including adipose tissue, liver, and muscle [Alhawiti et al, 2017]. [Szpigel et al, 2018] found that alterations in endogenous lipid metabolism and the plasma lipid environment in individuals with type 2 diabetes orientate monocytes towards a proinflammatory state through an ER stress–*TXNIP*–inflammasome pathway. Mutations in *TXNIP* were first associated with dyslipidemia in the HcB-19/Dem (HcB-19) mutant mouse with features resembling familial combined dyslipidemia including hypercholesterolemia, hypertriglyceridemia, and higher levels of triglyceride-rich lipoproteins [Bodnar JS, et al 2002].[Li, A, et al, 2023] highlighted that the potential role of *TXNIP* in cardiac dysfunction is induced by long-term high fat diet, where chronic exposure to dietary lipids is known to promote lipid deposition in the heart, which is the key factor for cardiac dysfunction. *TXNIP* deficiency increased cardiac HDL-C content compared with WT mice fed either with no diet or with high fat diet (HFD), suggesting that long-term HFD induced notable cardiac lipid deposition, which was remarkably reduced by *TXNIP* gene KO [Li, A, et al, 2023].

*PFKFB2* has been defined as the kidney isoform and also the cardiac isoform of the phosphofructokinase-2 enzyme (PFK-2), which is involved in the regulation of glycolysis and glucose metabolism [Harold, KM et al, 2024] [Lee M, et al, 2020].It is a critical glycolytic regulator responsible for upregulation of glycolysis in response to insulin and andrenergic signaling and its disruption impacts fatty acid oxidation [Harold KM et, 2024]. As a protein, it regulates fructose-2,6-biphosphate in the heart, while a related enzyme encoded by a different gene regulates fructose-2,6-biphosphate in the liver and muscle [NCBI, Gene database]. Impaired glycolysis due to disruption in *PFKFB2* expression can result in lipid accumulation, as the balance between glycolysis and lipogenesis is disrupted, possibly leading to increased levels of circulating free fatty acids and triglycerides, further aggravating insulin resistance [Gopalan C, et al,2022].

Both *PFKFB2* and *TXNIP* are associated with a group of metabolites in the lipid and amino acid metabolic pathways that are linked to HDL-C, LDL-C, and triglycerides [Yousri et al,2023]. Both are in linked to the glycolysis pathway and *TXNIP* has an mQTL located in GLUT1 [Yousri et al, 2023] that has been shown to impact cardiac function as that of *PFKFB2* [Harold, KM et al, 2024]. Moreover, both genes were found to be associated with the kidney and cardiac functions, highlighting the epigenetic role of those genes in the pathways associated with diabetes complications.

## Conclusion

This study has investigated the mechanisms relating T2D and altered lipids through methylation pathways in a middle eastern Qatari cohort. It identified *TXNIP* and *PFKFB2* as key genes linking T2D with triglyceride and lipoprotein levels and found associations with their genomic scores. These genes are key regulators of glucose and lipid metabolism, and their dysregulation contributes to the pathogenesis of diabetes and elevated triglycerides. *TXNIP* also promotes oxidative stress, inflammation, and lipid dysregulation, while *PFKFB2* influences glycolysis and lipogenesis. Understanding the molecular mechanisms underlying these pathways provides valuable insights into potential therapeutic targets for managing diabetes and dyslipidemia.

## Data Availability

All data produced in the present work are contained in the manuscript.

## Supplementary Data

**Supplementary Tables ST1-ST11:** Statistical results of GRS associations with their respective traits and stratified on gender and age.

**Supplementary Table ST12**: Statistical characteristics of traits.

**Supplementary Table ST13**: correlation of traits with age and BMI.

**Supplementary Table ST14**: Metabolites associated with lipid traits (adopted from [Yousri et al, 2023].

## Acknowledgments

This work was made possible by PPM2 grant #PPM2-0226–170020 from the Qatar National Research Fund (QNRF) and Qatar Precision and Health Institute(QPHI)-Qatar Genome Program (QGP) (members of Qatar Foundation). This work was also made possible by NPRP-S11 grant #NPRP11S-0114–180299 from the Qatar National Research Fund (a member of Qatar Foundation). The findings herein reflect the work and are solely the responsibility of the authors. We would like to acknowledge Qatar Precision and Health Institute – Qatar Bio Bank team for their support, and biomedical research at Weill Cornell Medicine-Qatar.

## Notes

### Competing Interest Statement

Authors have no competing interests, and the following declaration is for Ulrike Peters: U Peters was a consultant with AbbVie and her family is holding individual stocks for several companies or corporates.

### Author Declarations

IRB of Qatar Bio Bank gave ethical approval for this work. IRB number: QBB_IRB_E-2020-QF-QBB-RES-ACC-0104-0109

