## Supplementary Tables ST1-ST14 for "Epigenetic pathways and risk factors for Type 2 Diabetes in a Middle Eastern Cohort"

### Supplementary Table ST1: T2D GRS results

| Stratification | DF | beta | se | t-value | p-value | R2 | se_R2 | Lower-CI | Upper-CI |
|---|---|---|---|---|---|---|---|---|---|
| All | 10963 | 0.13 | 0.01 | 13.95 | 7.62E-44 | 0.02 | 0.002 | 0.01 | 0.02 |
| Males | 4762 | 0.16 | 0.01 | 11.20 | 9.44E-29 | 0.03 | 0.005 | 0.02 | 0.03 |
| Females | 6199 | 0.12 | 0.01 | 9.55 | 1.74E-21 | 0.01 | 0.003 | 0.01 | 0.02 |
| age <50 | 8558 | 0.02 | 0.01 | 1.80 | 7.11E-02 | 0.00 | 0.000 | 0.00 | 0.00 |
| age: 50-60 | 1556 | 0.20 | 0.02 | 7.85 | 7.56E-15 | 0.04 | 0.009 | 0.02 | 0.06 |
| age :60-70 | 666 | 0.14 | 0.04 | 3.66 | 2.75E-04 | 0.02 | 0.010 | 0.00 | 0.04 |
| age>70 | 177 | 0.14 | 0.07 | 1.90 | 5.92E-02 | 0.01 | 0.017 | -0.02 | 0.05 |

### Supplementary Table ST2 : LDL GRS - M1 (PT)

| | Beta | SE | Lower CI | Upper CI | t value | P | rsquare | se_r2 | r2.ci.lower | r2.ci.upper | n.obs |
|---|---|---|---|---|---|---|---|---|---|---|---|
| All | 0.26 | 0.01 | 0.25 | 0.28 | 28.49 | 3.26E-172 | 0.07 | 0.00 | 0.06 | 0.08 | 10879 |
| Males | 0.29 | 0.01 | 0.27 | 0.32 | 20.77 | 9.77E-92 | 0.08 | 0.01 | 0.07 | 0.10 | 4705 |
| Females | 0.27 | 0.01 | 0.24 | 0.29 | 21.99 | 3.02E-103 | 0.07 | 0.01 | 0.06 | 0.09 | 6174 |
| age < 50 | 0.28 | 0.01 | 0.26 | 0.30 | 27.19 | 3.50E-156 | 0.08 | 0.01 | 0.07 | 0.09 | 8495 |
| age 50-60 | 0.25 | 0.02 | 0.20 | 0.30 | 10.08 | 3.35E-23 | 0.06 | 0.01 | 0.04 | 0.09 | 1543 |
| age 60-70 | 0.23 | 0.04 | 0.16 | 0.30 | 6.35 | 3.98E-10 | 0.06 | 0.02 | 0.02 | 0.09 | 662 |
| age 70-80 | 0.23 | 0.08 | 0.08 | 0.38 | 3.04 | 2.79E-03 | 0.06 | 0.03 | -0.01 | 0.13 | 157 |
| age > 80 | -0.08 | 0.30 | -0.66 | 0.51 | -0.25 | 8.01E-01 | 0.00 | 0.02 | -0.04 | 0.05 | 22 |
| T2D | 0.23 | 0.02 | 0.19 | 0.27 | 11.55 | 5.30E-30 | 0.06 | 0.01 | 0.04 | 0.08 | 2195 |
| controls | 0.28 | 0.01 | 0.26 | 0.30 | 26.86 | 8.76E-153 | 0.08 | 0.01 | 0.07 | 0.09 | 8684 |

### Supplementary Table ST3: LDL GRS - M2 (PRSCS)

| | Beta | SE | Lower CI | Upper CI | t value | P | rsquare | se_r2 | r2.ci.lower | r2.ci.upper | n.obs |
|---|---|---|---|---|---|---|---|---|---|---|---|
| All | 0.27 | 0.01 | 0.26 | 0.29 | 29.64 | 9.05E-186 | 0.07 | 0.00 | 0.07 | 0.08 | 10879 |
| Males | 0.30 | 0.01 | 0.27 | 0.33 | 21.54 | 3.49E-98 | 0.09 | 0.01 | 0.07 | 0.11 | 4705 |
| Females | 0.28 | 0.01 | 0.26 | 0.30 | 22.95 | 6.38E-112 | 0.08 | 0.01 | 0.07 | 0.09 | 6174 |
| age < 50 | 0.30 | 0.01 | 0.28 | 0.32 | 28.79 | 5.79E-174 | 0.09 | 0.01 | 0.08 | 0.10 | 8495 |
| age 50-60 | 0.26 | 0.02 | 0.21 | 0.30 | 10.79 | 3.29E-26 | 0.07 | 0.01 | 0.05 | 0.09 | 1543 |
| age 60-70 | 0.21 | 0.04 | 0.14 | 0.29 | 5.81 | 9.76E-09 | 0.05 | 0.02 | 0.02 | 0.08 | 662 |
| age 70-80 | 0.14 | 0.08 | -0.01 | 0.29 | 1.79 | 7.56E-02 | 0.02 | 0.02 | -0.02 | 0.06 | 157 |
| age > 80 | 0.17 | 0.21 | -0.24 | 0.57 | 0.82 | 4.22E-01 | 0.03 | 0.06 | -0.10 | 0.16 | 22 |
| T2D | 0.25 | 0.02 | 0.21 | 0.29 | 12.23 | 2.68E-33 | 0.06 | 0.01 | 0.04 | 0.08 | 2195 |
| controls | 0.29 | 0.01 | 0.27 | 0.31 | 28.15 | 5.89E-167 | 0.08 | 0.01 | 0.07 | 0.09 | 8684 |

### Supplementary Table ST4: TC GRS - M1 (PT)

| | Beta | SE | Lower_CI | Upper_CI | t_value | P_value | rsquare | se_r2 | r2.ci.lower | r2.ci.upper | n.obs |
|---|---|---|---|---|---|---|---|---|---|---|---|
| All | 0.28 | 0.01 | 0.26 | 0.30 | 30.82 | 4.13E-200 | 0.08 | 0.00 | 0.07 | 0.09 | 10956 |
| Males | 0.32 | 0.01 | 0.29 | 0.35 | 23.03 | 2.24E-111 | 0.10 | 0.01 | 0.08 | 0.12 | 4761 |
| Females | 0.28 | 0.01 | 0.26 | 0.30 | 23.27 | 7.35E-115 | 0.08 | 0.01 | 0.07 | 0.09 | 6195 |
| age < 50 | 0.31 | 0.01 | 0.29 | 0.33 | 30.29 | 1.77E-191 | 0.10 | 0.01 | 0.08 | 0.11 | 8553 |
| age 50-60 | 0.25 | 0.02 | 0.21 | 0.30 | 10.57 | 2.86E-25 | 0.07 | 0.01 | 0.04 | 0.09 | 1557 |
| age 60-70 | 0.23 | 0.04 | 0.16 | 0.30 | 6.55 | 1.18E-10 | 0.06 | 0.02 | 0.03 | 0.10 | 667 |
| age 70-80 | 0.24 | 0.08 | 0.09 | 0.39 | 3.14 | 2.00E-03 | 0.06 | 0.04 | -0.01 | 0.13 | 157 |
| age > 80 | -0.01 | 0.27 | -0.55 | 0.52 | -0.05 | 9.64E-01 | 0.00 | 0.00 | -0.01 | 0.01 | 22 |
| T2D | 0.25 | 0.02 | 0.21 | 0.29 | 12.45 | 1.99E-34 | 0.07 | 0.01 | 0.05 | 0.08 | 2227 |
| controls | 0.31 | 0.01 | 0.29 | 0.33 | 30.02 | 1.68E-188 | 0.09 | 0.01 | 0.08 | 0.11 | 8729 |

### Supplementary Table ST5: TC GRS - M2 (PRSCS)

| | Beta | SE | Lower_CI | Upper_CI | t_value | P_value | rsquare | se_r2 | r2.ci.lower | r2.ci.upper | n.obs |
|---|---|---|---|---|---|---|---|---|---|---|---|
| All | 0.29 | 0.01 | 0.27 | 0.30 | 31.17 | 2.24E-204 | 0.08 | 0.01 | 0.07 | 0.09 | 10956 |
| Males | 0.32 | 0.01 | 0.30 | 0.35 | 23.70 | 1.96E-117 | 0.11 | 0.01 | 0.09 | 0.12 | 4761 |
| Females | 0.28 | 0.01 | 0.26 | 0.31 | 23.10 | 2.69E-113 | 0.08 | 0.01 | 0.07 | 0.09 | 6195 |
| age < 50 | 0.32 | 0.01 | 0.30 | 0.34 | 30.71 | 1.54E-196 | 0.10 | 0.01 | 0.09 | 0.11 | 8553 |
| age 50-60 | 0.26 | 0.02 | 0.21 | 0.31 | 10.84 | 1.94E-26 | 0.07 | 0.01 | 0.05 | 0.09 | 1557 |
| age 60-70 | 0.21 | 0.04 | 0.14 | 0.28 | 5.84 | 7.98E-09 | 0.05 | 0.02 | 0.02 | 0.08 | 667 |
| age 70-80 | 0.18 | 0.07 | 0.04 | 0.33 | 2.50 | 1.33E-02 | 0.04 | 0.03 | -0.02 | 0.10 | 157 |
| age > 80 | -0.01 | 0.22 | -0.45 | 0.43 | -0.04 | 9.67E-01 | 0.00 | 0.00 | -0.01 | 0.01 | 22 |
| T2D | 0.25 | 0.02 | 0.21 | 0.29 | 12.56 | 5.22E-35 | 0.07 | 0.01 | 0.05 | 0.09 | 2227 |
| controls | 0.31 | 0.01 | 0.29 | 0.33 | 30.39 | 6.96E-193 | 0.10 | 0.01 | 0.08 | 0.11 | 8729 |

### Supplementary Table ST6: HDL GRS - M1 (PT)

| | Beta | SE | Lower_CI | Upper_CI | t_value | P_value | rsquare | se_r2 | r2.ci.lower | r2.ci.upper | n.obs |
|---|---|---|---|---|---|---|---|---|---|---|---|
| All | 0.26 | 0.01 | 0.25 | 0.28 | 28.63 | 6.09E-174 | 0.07 | 0.00 | 0.06 | 0.08 | 10959 |
| Males | 0.30 | 0.01 | 0.27 | 0.33 | 21.52 | 4.10E-98 | 0.09 | 0.01 | 0.07 | 0.10 | 4762 |
| Females | 0.28 | 0.01 | 0.25 | 0.30 | 22.95 | 5.80E-112 | 0.08 | 0.01 | 0.07 | 0.09 | 6197 |
| age < 50 | 0.25 | 0.01 | 0.23 | 0.27 | 23.71 | 2.20E-120 | 0.06 | 0.01 | 0.05 | 0.07 | 8556 |
| age 50-60 | 0.32 | 0.02 | 0.27 | 0.37 | 13.29 | 2.97E-38 | 0.10 | 0.01 | 0.07 | 0.13 | 1556 |
| age 60-70 | 0.31 | 0.04 | 0.24 | 0.39 | 8.19 | 1.33E-15 | 0.09 | 0.02 | 0.05 | 0.13 | 668 |
| age 70-80 | 0.25 | 0.07 | 0.11 | 0.40 | 3.41 | 8.41E-04 | 0.07 | 0.04 | -0.01 | 0.15 | 157 |
| age > 80 | -0.05 | 0.21 | -0.46 | 0.36 | -0.23 | 8.21E-01 | 0.00 | 0.02 | -0.04 | 0.04 | 22 |
| T2D | 0.29 | 0.02 | 0.25 | 0.33 | 14.40 | 5.10E-45 | 0.09 | 0.01 | 0.06 | 0.11 | 2229 |
| controls | 0.25 | 0.01 | 0.23 | 0.27 | 23.80 | 2.51E-121 | 0.06 | 0.00 | 0.05 | 0.07 | 8730 |

### Supplementary Table ST7: HDL GRS - M2 (PRSCS)

| | Beta | SE | Lower_CI | Upper_CI | t_value | P_value | rsquare | se_r2 | r2.ci.lower | r2.ci.upper | n.obs |
|---|---|---|---|---|---|---|---|---|---|---|---|
| All | 0.25 | 0.01 | 0.24 | 0.27 | 27.50 | 4.76E-161 | 0.06 | 0.00 | 0.06 | 0.07 | 10959 |
| Males | 0.29 | 0.01 | 0.26 | 0.31 | 20.68 | 4.95E-91 | 0.08 | 0.01 | 0.07 | 0.10 | 4762 |
| Females | 0.27 | 0.01 | 0.25 | 0.29 | 21.96 | 5.15E-103 | 0.07 | 0.01 | 0.06 | 0.08 | 6197 |
| age < 50 | 0.24 | 0.01 | 0.22 | 0.26 | 22.75 | 3.00E-111 | 0.06 | 0.00 | 0.05 | 0.07 | 8556 |
| age 50-60 | 0.31 | 0.02 | 0.26 | 0.36 | 12.84 | 5.90E-36 | 0.10 | 0.01 | 0.07 | 0.12 | 1556 |
| age 60-70 | 0.34 | 0.04 | 0.27 | 0.41 | 9.29 | 2.07E-19 | 0.11 | 0.02 | 0.07 | 0.16 | 668 |
| age 70-80 | 0.28 | 0.08 | 0.12 | 0.43 | 3.48 | 6.47E-04 | 0.07 | 0.04 | 0.00 | 0.15 | 157 |
| age > 80 | -0.06 | 0.18 | -0.40 | 0.28 | -0.34 | 7.37E-01 | 0.01 | 0.03 | -0.05 | 0.06 | 22 |
| T2D | 0.29 | 0.02 | 0.25 | 0.33 | 14.03 | 6.86E-43 | 0.08 | 0.01 | 0.06 | 0.10 | 2229 |
| controls | 0.24 | 0.01 | 0.22 | 0.26 | 22.92 | 5.43E-113 | 0.06 | 0.00 | 0.05 | 0.07 | 8730 |

### Supplementary Table ST8: TG GRS - M1 (PT)

| | Beta | SE | Lower_CI | Upper_CI | t_value | P_value | rsquare | se_r2 | r2.ci.lower | r2.ci.upper | n.obs |
|---|---|---|---|---|---|---|---|---|---|---|---|
| All | 0.20 | 0.01 | 0.18 | 0.22 | 21.45 | 5.66E-100 | 0.04 | 0.00 | 0.03 | 0.05 | 10961 |
| Males | 0.22 | 0.01 | 0.19 | 0.25 | 14.98 | 1.31E-49 | 0.05 | 0.01 | 0.03 | 0.06 | 4762 |
| Females | 0.21 | 0.01 | 0.19 | 0.23 | 17.22 | 6.03E-65 | 0.05 | 0.01 | 0.04 | 0.06 | 6199 |
| age < 50 | 0.21 | 0.01 | 0.19 | 0.23 | 19.52 | 4.84E-83 | 0.04 | 0.00 | 0.03 | 0.05 | 8556 |
| age 50-60 | 0.21 | 0.02 | 0.16 | 0.26 | 8.68 | 9.91E-18 | 0.05 | 0.01 | 0.03 | 0.07 | 1558 |
| age 60-70 | 0.21 | 0.04 | 0.14 | 0.28 | 5.62 | 2.84E-08 | 0.05 | 0.02 | 0.01 | 0.08 | 668 |
| age 70-80 | 0.23 | 0.08 | 0.08 | 0.38 | 2.92 | 4.00E-03 | 0.05 | 0.03 | -0.01 | 0.12 | 157 |
| age > 80 | -0.20 | 0.24 | -0.67 | 0.26 | -0.85 | 4.03E-01 | 0.04 | 0.07 | -0.10 | 0.17 | 22 |
| T2D | 0.18 | 0.02 | 0.14 | 0.22 | 9.01 | 4.22E-19 | 0.04 | 0.01 | 0.02 | 0.05 | 2229 |
| controls | 0.21 | 0.01 | 0.19 | 0.23 | 19.76 | 4.72E-85 | 0.04 | 0.00 | 0.03 | 0.05 | 8732 |

### Supplementary Table ST9: TG GRS - M2 (PRSCS)

| | Beta | SE | Lower_CI | Upper_CI | t_value | P_value | rsquare | se_r2 | r2.ci.lower | r2.ci.upper | n.obs |
|---|---|---|---|---|---|---|---|---|---|---|---|
| All | 0.24 | 0.01 | 0.22 | 0.26 | 25.57 | 4.45E-140 | 0.06 | 0.00 | 0.05 | 0.06 | 10961 |
| Males | 0.23 | 0.01 | 0.21 | 0.26 | 16.49 | 1.94E-59 | 0.05 | 0.01 | 0.04 | 0.07 | 4762 |
| Females | 0.26 | 0.01 | 0.24 | 0.29 | 21.51 | 5.02E-99 | 0.07 | 0.01 | 0.06 | 0.08 | 6199 |
| age < 50 | 0.24 | 0.01 | 0.22 | 0.26 | 22.50 | 6.26E-109 | 0.06 | 0.00 | 0.05 | 0.07 | 8556 |
| age 50-60 | 0.28 | 0.02 | 0.23 | 0.33 | 11.67 | 3.22E-30 | 0.08 | 0.01 | 0.05 | 0.11 | 1558 |
| age 60-70 | 0.28 | 0.04 | 0.21 | 0.35 | 7.54 | 1.58E-13 | 0.08 | 0.02 | 0.04 | 0.12 | 668 |
| age 70-80 | 0.25 | 0.07 | 0.11 | 0.39 | 3.48 | 6.56E-04 | 0.07 | 0.04 | 0.00 | 0.15 | 157 |
| age > 80 | -0.19 | 0.20 | -0.58 | 0.19 | -0.97 | 3.44E-01 | 0.04 | 0.07 | -0.11 | 0.20 | 22 |
| T2D | 0.25 | 0.02 | 0.21 | 0.29 | 12.60 | 3.34E-35 | 0.07 | 0.01 | 0.05 | 0.09 | 2229 |
| controls | 0.24 | 0.01 | 0.22 | 0.26 | 22.51 | 4.31E-109 | 0.05 | 0.00 | 0.05 | 0.06 | 8732 |

### Supplementary Table ST10: Non-HDL GRS - M1 (PT)

| | Beta | SE | Lower_CI | Upper_CI | t_value | P_value | rsquare | se_r2 | r2.ci.lower | r2.ci.upper | n.obs |
|---|---|---|---|---|---|---|---|---|---|---|---|
| All | 0.21 | 0.01 | 0.19 | 0.22 | 22.10 | 6.29E-106 | 0.04 | 0.004 | 0.04 | 0.05 | 10954 |
| Males | 0.23 | 0.01 | 0.20 | 0.25 | 15.86 | 2.96E-55 | 0.05 | 0.006 | 0.04 | 0.06 | 4761 |
| Females | 0.22 | 0.01 | 0.19 | 0.24 | 17.51 | 4.65E-67 | 0.05 | 0.005 | 0.04 | 0.06 | 6193 |
| age < 50 | 0.21 | 0.01 | 0.19 | 0.23 | 19.79 | 2.68E-85 | 0.04 | 0.004 | 0.04 | 0.05 | 8553 |
| age 50-60 | 0.22 | 0.02 | 0.17 | 0.26 | 8.86 | 2.17E-18 | 0.05 | 0.011 | 0.03 | 0.07 | 1555 |
| age 60-70 | 0.16 | 0.04 | 0.08 | 0.23 | 4.13 | 4.14E-05 | 0.02 | 0.012 | 0.00 | 0.05 | 667 |
| age 70-80 | 0.14 | 0.08 | -0.02 | 0.29 | 1.74 | 8.30E-02 | 0.02 | 0.021 | -0.02 | 0.06 | 157 |
| age > 80 | -0.24 | 0.27 | -0.77 | 0.30 | -0.86 | 4.00E-01 | 0.04 | 0.066 | -0.10 | 0.17 | 22 |
| T2D | 0.19 | 0.02 | 0.15 | 0.23 | 9.04 | 3.23E-19 | 0.04 | 0.008 | 0.02 | 0.05 | 2227 |
| controls | 0.21 | 0.01 | 0.19 | 0.23 | 19.84 | 9.77E-86 | 0.04 | 0.004 | 0.03 | 0.05 | 8727 |

### Supplementary Table ST11: Non-HDL GRS - M2 (PRSCS)

| | Beta | SE | Lower_CI | Upper_CI | t_value | p-value | rsquare | se_r2 | r2.ci.lower | r2.ci.upper | n.obs |
|---|---|---|---|---|---|---|---|---|---|---|---|
| All | 0.24 | 0.01 | 0.22 | 0.26 | 25.89 | 1.52E-143 | 0.06 | 0.00 | 0.05 | 0.07 | 10954 |
| Males | 0.26 | 0.01 | 0.24 | 0.29 | 18.75 | 1.02E-75 | 0.07 | 0.01 | 0.05 | 0.08 | 4761 |
| Females | 0.25 | 0.01 | 0.22 | 0.27 | 20.34 | 4.66E-89 | 0.06 | 0.01 | 0.05 | 0.07 | 6193 |
| age < 50 | 0.25 | 0.01 | 0.23 | 0.27 | 23.23 | 7.92E-116 | 0.06 | 0.00 | 0.05 | 0.07 | 8553 |
| age 50-60 | 0.24 | 0.02 | 0.20 | 0.29 | 10.20 | 1.05E-23 | 0.06 | 0.01 | 0.04 | 0.09 | 1555 |
| age 60-70 | 0.20 | 0.04 | 0.12 | 0.27 | 5.33 | 1.37E-07 | 0.04 | 0.01 | 0.01 | 0.07 | 667 |
| age 70-80 | 0.15 | 0.08 | 0.00 | 0.30 | 2.02 | 4.52E-02 | 0.03 | 0.02 | -0.02 | 0.07 | 157 |
| age > 80 | 0.08 | 0.24 | -0.38 | 0.54 | 0.34 | 7.37E-01 | 0.01 | 0.03 | -0.05 | 0.06 | 22 |
| T2D | 0.24 | 0.02 | 0.20 | 0.28 | 11.90 | 1.01E-31 | 0.06 | 0.01 | 0.04 | 0.08 | 2227 |
| control | 0.24 | 0.01 | 0.22 | 0.26 | 22.62 | 4.03E-110 | 0.06 | 0.00 | 0.05 | 0.06 | 8727 |

### Supplementary Table ST12: Statistics of clinical traits

| Trait | N | mean | sd | Female | Age.mean | Age.sd | T2D.Case | n.SNP (M1) | n.SNP (M2) |
|---|---|---|---|---|---|---|---|---|---|
| HDL-C | 10959 | 55.2 | 15.2 | 57% | 38.7 | 13.1 | 20% | 8621 | 1147143 |
| LDL-C | 10879 | 115.4 | 35.5 | 57% | 38.7 | 13.1 | 20% | 7508 | 1147143 |
| TG | 10961 | 4.6 | 0.5 | 57% | 38.7 | 13.1 | 20% | 7673 | 1147143 |
| nonHDL-C | 10954 | 137.2 | 40.5 | 57% | 38.7 | 13.1 | 20% | 6731 | 1147143 |
| TC | 10956 | 192.3 | 38.8 | 57% | 38.7 | 13.1 | 20% | 8668 | 1147143 |

### Supplementary Table ST13: Correlation of lipid traits with Age and BMI

| | corr Age and lipids | | corr BMI and lipids | |
|---|---|---|---|---|
| | cor | pval | cor | pval |
| LDL | 0.25 | 1.14E-15 | 0.13 | 3.46E-05 |
| TC | 0.29 | 7.52E-22 | 0.14 | 5.07E-06 |
| HDL | -0.07 | 2.92E-02 | -0.12 | 1.37E-04 |
| TG | 0.24 | 1.33E-15 | 0.16 | 1.59E-07 |
| nonHDL | 0.31 | 3.74E-24 | 0.18 | 8.18E-09 |

### Supplementary Table ST14: Metabolites associated with lipids from [Yousri et al, 2023]

| Metabolites | Discovery p-value | Replication p-value | Phenotype |
|---|---|---|---|
| 1-(1-enyl-palmitoyl)-2-oleoyl-GPC (P-16:0/18:1)* | 1.3784E-104 | 1.17E-72 | HDL-C |
| 1-(1-enyl-palmitoyl)-2-palmitoleoyl-GPC (P-16:0/16:1)* | 2.64911E-54 | 4.85E-46 | HDL-C |
| 1-(1-enyl-palmitoyl)-2-palmitoyl-GPC (P-16:0/16:0)* | 9.80245E-42 | 4.76E-33 | HDL-C |
| 1-(1-enyl-palmitoyl)-GPC (P-16:0)* | 3.38775E-43 | 1.79E-38 | HDL-C |
| glycosyl ceramide (d18:2/24:1, d18:1/24:2)* | 2.51849E-16 | 1.20E-08 | LDL-C |
| glycosyl-N-palmitoyl-sphingosine (d18:1/16:0) | 1.32216E-58 | 4.64E-40 | LDL-C |
| lactosyl-N-palmitoyl-sphingosine (d18:1/16:0) | 3.54185E-43 | 3.06E-26 | LDL-C |
| 3-methyl-2-oxobutyrate | 3.93E-14 | 3.14E-03 | BMI |
| 1-carboxyethylphenylalanine | 1.4214E-34 | 4.62E-21 | TRI |
| 1-carboxyethylvaline | 2.31638E-12 | 3.96E-11 | TRI |
| 1-palmitoyl-2-arachidonoyl-GPE (16:0/20:4)* | 1.04716E-76 | 2.98E-70 | TRI |
| 1-palmitoyl-2-oleoyl-GPE (16:0/18:1) | 6.4711E-100 | 1.40E-80 | TRI |
| 3-methyl-2-oxovalerate | 1.13319E-14 | 9.77E-09 | TRI |
| alanine | 9.34006E-13 | 9.34E-13 | TRI |
| X - 14056 | 1.64136E-16 | 1.64E-16 | TRI |
| X - 19438 | 1.08632E-12 | 1.09E-12 | TRI |
